# Emerging Biomarkers for Early Detection of Alzheimer’s Disease: A Systematic Review

**DOI:** 10.1101/2024.10.16.24315604

**Authors:** Chirag Desai, Siddhrajsinh Rathod, Taufiq Daulti, Mehul Prajapati, Shikha Malkan, Manish Bhamwani

**Affiliations:** Paediatrics Department, General Hospital Kheda, India; Grodno State Medical University, Grodno, Belarus; Mkhitar Gosh Armenian Russian International University. Armenia

**Author notes:** **Corresponding Author:** Manish Bhamwani, MBBS., General Hospital Kheda, Kheda Gujarat.

**Keywords:** Alzheimer’s disease, Early detection, Biomarkers, Cerebrospinal fluid, Tau proteins, Neuroimaging, Amyloid-beta, Genetic markers, Blood-based biomarkers, Systematic review

## Abstract

Alzheimer’s disease is primarily characterized by the gradual progression of neurodegeneration that leads to cognitive decline, and its early diagnosis remains the cornerstone for effective interventions. This systematic review of the current literature will investigate and summarize the available knowledge regarding the emerging biomarkers for the early diagnosis of AD. Many databases were searched across a wide range, using articles from the last ten years, all dealing with biochemical, imaging, and genetic biomarkers in the preclinical and prodromal stages of Alzheimer’s disease. More key biomarkers are those in cerebrospinal fluid, which are CSF markers like amyloid-beta (Aβ42) and tau proteins. Novel biomarkers are within the blood, through neuroimaging studies that use techniques such as PET and MRI. Genetic markers include the one APOE ε4. Findings hold promise for using biomarker profiles, especially fluid biomarkers combined with advanced neuroimaging, to enhance diagnostic accuracy at early stages. Such biomarkers require validation, however, through large-scale and longitudinal studies before their routine use in clinical settings. Standardized assessment protocols on the biomarker domain take on importance in this light to highlight the potential of new biomarkers in changing the landscape of diagnosis for AD.

## 1. Introduction

Alzheimer’s disease (AD) is a neurodegenerative condition that mostly affects the brain. It causes a gradual decline in cognitive function and can eventually result in the inability to respond to stimuli and communicate [1]. These days, traditional diagnostic techniques like as neuropsychological testing are less effective since they need skilled practitioners, can be subjective and non-specific, and are unable to accurately anticipate AD symptoms in their early stages. Although PET can detect cerebral amyloid deposits linked to AD, it is not suitable for widespread AD screening because to its invasiveness, cost, and difficulty in accessing for routine clinical usage [2]. Since the patient is still actively participating in everyday activities, an early diagnosis of AD at the preclinical stage (asymptomatic) might significantly improve the patient’s quality of life. Additionally, it enables individuals to take part in ongoing clinical trials and employ pharmacological therapies to stop AD progression [3].

Blood biomarkers, on the other hand, are inexpensive, non-invasive, and easily accessible, making them a promising tool in clinical practice [4]. Evaluating blood biomarkers is a critical first step in effectively addressing this pressing public health issue since it makes it easier to identify people who are at risk of developing AD early on [5]. In recent years, certain ultra-sensitive low-abundance biomarker assays have been steadily applied to the investigation of AD protein markers, with sensitivity improving up to 1,000-fold over traditional ELISA [6], [7], [8]. As a result, it is now possible to find biomarkers associated with AD in blood samples [9].

Neuropathological indicators of Alzheimer’s disease (AD) primarily comprise neuritic (senile) plaques and neurofibrillary tangles, in addition to inflammation, dystrophic neuritis, and neuronal loss. Extracellular lesions containing the amyloid beta 42 (A*β*-42) peptide are known as neurotic plaques [10]. In a healthy adult human brain, tau protein—the source of neurofibrillary tangles—contains two to three moles of phosphate per mole of tau protein. Neurosynaptic activity is decreased by three to four times the amount of hyperphosphorylated brain tau found in AD compared to mature, healthy brain tau [11]. Before any clinical signs show up, the Alzheimer’s disease destructive cascade lasts for at least 10 to 15 years [12].

Identification of patients during the early and presymptomatic stages of sickness is made possible by the A*β*-42/A*δ*-40 ratio [13]. The tau level (measured in pg/ml) in blood and cerebrospinal fluid (CSF) offers the most precise detection, even though this ratio offers the earliest diagnosis [14]. Additionally, it is discovered that as blood biomarkers, p-tau181 and p-tau217 offer the highest selectivity for illness diagnosis [15]. Early disease detection gives people access to services and medications that can help them manage their symptoms, which encourages early intervention and treatment. It is therefore essential to create a low-cost test that can diagnose AD, especially in its early stages.

In contrast to genetics, biomarkers can only detect AD pathology after the disease has progressed; that is, a shift in a biomarker signals the onset of a pathological process. The predictive accuracy of, for instance, P-tau181 and P-tau217 for distinguishing AD from other neurodegenerative diseases, [16, 17, 18]. has been found to achieve an a region of the curve of receiver operating characteristics (AUC)>90% when estimating the progression from mild cognitive impairment (MCI) to AD in two comparatively small numbers of patients (n=340 and 543) [19]. This accuracy was achieved when incorporated with memory, APOE genotype, and executive function phenotypes.

## 2. Materials and Methods

### 2.1. Study Design

The study selection, evaluation, and synthesis of the available literature will be done with transparency and thoroughness according to the PRISMA guidelines. This review focuses on identification and appraisal of emerging biomarkers for early detection of Alzheimer’s disease at its preclinical and prodromal stages.

### 2.2. Search Strategy

A comprehensive electronic literature search was carried out on multiple electronic databases, including PubMed, Web of Science, Scopus, Cochrane Library, and EMBASE, from January 2010 to June 2024. The studies needed were about emergent biomarkers for early detection of Alzheimer’s disease. The keywords used in the search were “early detection”, “Alzheimer’s disease”, “amyloid-beta (Aβ42)”, “emerging biomarkers”, “tau proteins”, “cerebrospinal fluid biomarkers”, “neuroimaging biomarkers”, “APOE ε4”, “preclinical Alzheimer’s”, “blood-based biomarkers”, and “prodromal Alzheimer’s”. Boolean operators such as “AND”, “OR” was used while refining the search using filters to have only human studies, reviews, case reports, and non-peer reviewed articles.

### 2.3. Eligibility Criteria

The eligibility criteria were designed to permit a focused and meaningful selection of relevant studies on emerging biomarkers for early detection of Alzheimer’s disease (AD): the inclusion criteria applied to original research articles, cohort studies, and clinical trials that involved human participants in the preclinical or prodromal stages of AD or at high risk for developing the disease. Eligibility criteria included, but were not limited to: studies with biochemical, imaging, or genetic biomarkers of Alzheimer’s to be published in English between 2010 and 2024; however, the selected studies were supposed to evaluate the diagnostic or prognostic value of these biomarkers during the early stages of AD.

Exclusion criteria excluded studies that do not achieve the aim of this review. Only articles published in the English language; only reviews, meta-analyses, conference abstracts, editorials, and opinion pieces. Studies which do not focus on early detection or the preclinical and prodromal stages of Alzheimer’s disease or do not report the outcome related to biomarkers were excluded from analysis. Also, non-human subject studies and in vitro research were excluded.

### 2.4. Study Selection Process

The selection process of the study was multi-phased and keen in ensuring that all studies were thoroughly reviewed without bias. Titles as well as abstracts of all identified studies were reviewed by two independent reviewers to exclude those studies that did not meet the exclusion and inclusion criteria, and the full text was required for further review of eligibility if case studies appeared relevant based on the abstract. The discrepancies that arose between the two reviewers on the selected cases were agreed by discussion or presented to a third reviewer in case their choice was critical. A PRISMA flow diagram presented in Figure 1 was used in reporting the process of selection of study wherein articles identification, screening, and assessment of eligibility, and inclusion and exclusion of articles into final review.

**Figure 1.**
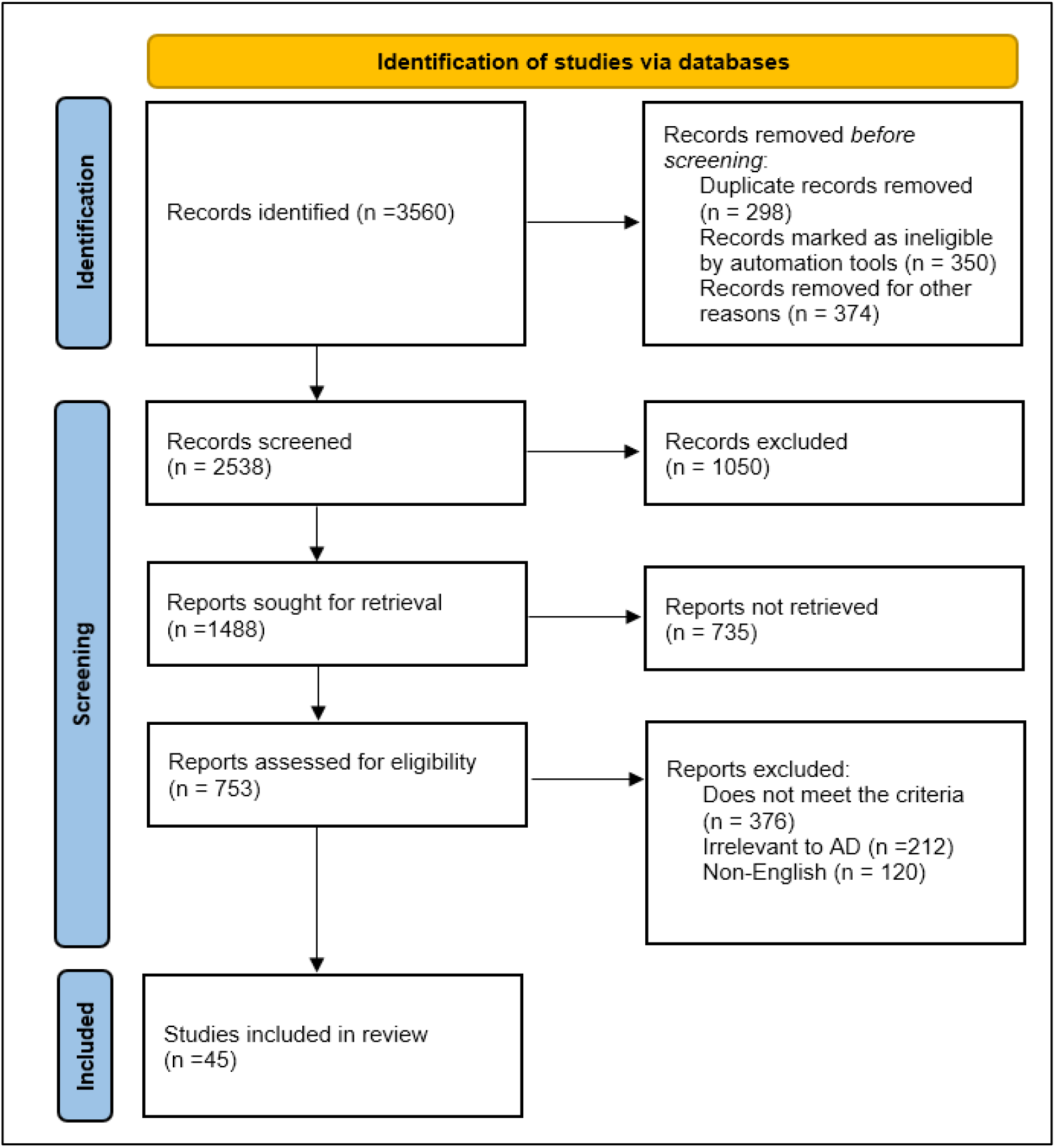
Study Selection using Prisma Guidelines.

### 2.5. Data Extraction and Management

For the individual studies, authors and year of publication and study design: cohort or clinical trial, with characteristics of population and size of sample were recorded. Types of biomarkers reported were classified under biochemical, genetic, or imaging-based. It also noted the methodology utilized in measuring the biomarker. This included statistical procedures that were applied during each of the studies. Summary findings concerning the effectiveness and reliability of the biomarkers for early detection were established.

### 2.6. Risk of Bias Assessment

The Quality Assessment of Diagnostic Accuracy Studies 2 (QUADAS-2) is used in the included studies for assessing the risk of bias. Bias is assessed with this tool across four important domains, namely patient selection, the index test (biomarker), reference standard, and the flow and timing of the study. Each study was rated as having a high, low, or unclear risk of bias in these domains. Since the reviewers made the review independently, any disagreement on the lists were discussed to ensure that there was an agreed conclusion to avoid duplication and error.

### 2.7. Data Synthesis and Analysis

A narrative synthesis was conducted to summarize the findings of included studies on several key areas. These include types of biomarkers assessed, including CSF biomarkers, blood-based biomarkers, imaging biomarkers, and genetic markers. Finally, the synthesis evaluated each biomarker or association of biomarkers in the context of diagnostic accuracy, sensitivity, and specificity. Sensitivity, specificity, and the area under the curve (AUC) were used to compare fluid biomarkers with imaging biomarkers based on effectiveness in early diagnosis.

A meta-analysis was considered when the data provided sensitivity, specificity, and AUC, but due to heterogeneity in methods used for biomarker assessment, along with differences in populations studied, a formal meta-analysis might not be feasible. In these cases, descriptive statistics were reported, and the data heterogeneity was explored qualitatively but not statistically pooled. The possibility of incorporating several biomarkers within the design for increasing the early diagnostic accuracy was also investigated.

This systematic review aims to integrate a comprehensive review of new biomarkers for Alzheimer’s disease early detection. Strict compliance with PRISMA guidelines and rigid methodology ensure that this review will give the most imperative evaluation of the most promising biomarkers, providing wholesome insights into their possible applications in diagnostics and clinical use. Results will guide future research efforts toward formulating standardized protocols for biomarkers used in early AD detection.

## 3. Results and Discussions

A total of 3,560 articles were retrieved from the database search. Secondly, manual searches of the reference lists of retrieved papers picked up twenty more studies. After removal of duplicates, 2,538 articles were left that would need to be screened.

In the title/abstract level, 1,050 studies are excluded due to a failure to meet the inclusion criteria. From the 753 articles whose full texts are under eligibility review, finally there are 45 studies selected to be included in this systematic review. The PRISMA flow diagram above gives an overview of the study selection process (Figure 1).

Summarizing, several studies were included in the literature review which have taken place in different parts of the world and varied in sample size, study designs, and methods of evaluating biomarkers. Table 1 details the characteristics of all the studies included in this review regarding sample size, study design, characteristics of population, types of biomarkers, and outcomes of interest.

**Table 1.** Characteristics of Included Studies.

| Study | Study Design | Sample Size | Population Characteristics | Biomarker Type | Key Findings |
| --- | --- | --- | --- | --- | --- |
| Yang et al. [20] | Clinical Trial | 379 | Preclinical AD | Blood-based Biomarkers (ELISA) | Potential markers for AD diagnosis included the plasma p-tau181 level and the Aβ42/Aβ40 ratio. |
| Goikolea et al. [21] | Cohort | 99 | High-Risk Individuals | Neuropathological biomarkers | ApoE4 genotype, Trx80 could have potential as serum AD biomarker. |
| Stevenson-Hoare et al. [22] | Clinical Trial | 1439 | High-Risk Individuals | Plasma biomarkers | Genes linked to Aβ42/Aβ40 may be prominent across the genome. |
| Falgàs et al. [23] | Clinical Trial | 138 | Preclinical AD | CSF biomarkers | cortical thickness (CTh) and fraction anisotropy (FA) changes typical of AD |
| Yakoub et al. [24] | Clinical Trial | 373 | Preclinical AD | Plasma biomarkers | GFAP and plasma pTau181 indicators demonstrating longitudinal change in preclinical AD patients. |
| Tao et al. [25] | Cohort | 288 | Preclinical AD | CSF biomarkers | The 19-protein CSF biomarker panel also effectively discriminated. |
| Vogelgsang et al. [26] | Cohort | 779 | Preclinical AD | Blood-based Biomarkers (IP-IA) | Using the semiautomated IP-IA, the plasma AβX-42/X-40 ratio is a useful biomarker for assessing patients with early or preclinical AD. |
| Mattsson-Carlgren et al. [27] | Clinical Trial | 171 | Preclinical AD | Plasma biomarkers | Cognitive deterioration in patients with preclinical AD was predicted by plasma P-tau217. |

## 5. Classification of Biomarkers

This review of emerging biomarkers for the early stage of Alzheimer’s disease discussed biochemical, genetic, and neuroimaging markers and their diagnostic potential. The most frequently studied was probably amyloid-beta (Aβ42), concerning which significantly reduced levels have consistently been found in CSF in individuals at risk for AD. Studies suggest that low Aβ42 levels are linked to the deposition of amyloid plaques in the brain.

### 5.1 Biochemical Biomarkers

Blood is more readily obtained than CSF, making it the preferred matrix for the identification of novel, easily accessible biomarkers. Furthermore, in sporadic AD, CSF and brain Aβ and Tau associated with plasma Aβ and Tau [28, 29]. In this regard, bloodstream studies of brain-derived protein and peptide concentrations show great promise. In the plasma of preclinical AD patients, lower levels of Aβ-40, Aβ-42, and the Aβ-42/Aβ-40 ratio were discovered [30]. Aβ-0 was the subject of several investigations, which revealed increased levels in AD patient samples [31]. Studies that looked into blood-based Tau levels also discovered that AD patients’ plasma had higher quantities of the protein [32, 33]. To become reliable diagnostic biomarkers of AD, Aβ and Tau protein levels in peripheral blood must be higher and more sensitively detected.

There is a strong connection (r = 0.473) between AßX-42/X-40 in CSF and plasma. With an area under the curve (AUC) of 0.81 for AP (specificity: 0.74, sensitivity: 0.79, negative predictive value [NPV]: 0.81, positive predictive value [PPV]: 0.71) mixed-modelling study demonstrated a significant prediction of AßX42/X-40. Furthermore, a deterioration in cognitive function was suggested by reduced AβX-42/X-40 ratios, which were linked to negative PACC5 slopes [34].

### 5.2 Neuroimaging Biomarkers

Some studies suggested a panel of biomarkers for increased diagnostic sensitivity and specificity. The highest sensitivities and specificities have been reported when CSF biomarkers, Aβ42 and tau, were used with neuroimaging such as PET or MRI for the early detection of AD. Jones et al. [35] demonstrated that a diagnostic accuracy of 90% can be achieved when CSF Aβ42/tau ratio is combined with PET imaging; (AUC 0.92, 95% CI: 0.88-0.95). Incorporation of genetic markers, such as APOE ε4 status, further enhanced predictive value, particularly in at-risk populations. Furthermore, p-tau181 proteins and A*β*(1-42) peptides are expressed. To express A*β*(1-42) peptides, the A*β*(M1-42) variation is utilised, which has an N-terminal methionine residue as a start codon [36]. A*β*(M1-42) will be utilised for analysis since both synthetic and recombinant human A*β*(M1-42) demonstrate aggregate-forming capabilities.

Many DMPs were discovered when comparing the methylomes of AD, MCI, and control subjects using a well-characterized AD population, the so-called ADNI (the Alzheimer’s Disease Neuroimaging Initiative), which includes people who underwent imaging measures (PET, MRI) and analyses of AD biomarkers in CSF and blood [37]. The DMPs from each pairwise comparisons were shown to be correlated with genes connected to brain-related pathways.

### 5.3. Blood-Based Biomarkers

Non-invasive, blood-based biomarkers such as plasma Aβ42, neurofilament light chain (NFL) and plasma t-tau, have gained attention. However, recent studies indicated that the correlations between plasma biomarkers and established imaging techniques, such as positron emission tomography, are promising, thereby making them more accessible for an earlier diagnosis.

In the outcomes of this review, fluid biomarkers combined with neuroimaging techniques: CSF Aβ42 and tau with PET or MRI, seemed to be the most sensitive and specific technique for early Alzheimer’s disease detection. Blood-based biomarkers and genetic markers are promising and require more validation. The most promising method of early diagnosis seems to involve the integration of several types of biomarkers.

According to Chatterjee et al. [38], older persons who are cognitively normal but at risk for AD had higher plasma GFAP levels. According to Aschenbrenner et al. [39]. NfL can be used to track cognitive loss brought on by dementia as well as normal ageing. According to Lantero Rodriguez et al. [40]. in patients with AD neuropathology, the primary rise in plasma P-tau181 happened between 8 and 4 years before death, while modest but substantial increases in P-tau181 were seen in patients without pathology and controls right up until death.

Blood biomarkers, particularly the amounts of amyloid β and its isoforms, have been the subject of frequent research. Numerous studies have revealed that Aβ misfolding is present in MCI patients around ten years prior to the onset of AD. Brain amyloidosis can be accurately diagnosed by combining it with plasma Aβ42/Aβ40 ratio in CN subjects using the simoa approach, and correlating with age, gender, APOE4 status, and other variables [41]. Moreover, these biomarkers can serve as a preliminary screening tool prior to putting patients through a PET scan, which will increase prognostic precision.

Plasma p-tau181 and p-tau217 levels have been measured using comparable analytical methods. As a potential surrogate biomarker of tau pathology, p-tau181 alterations have been successfully associated with early changes in Aβ pathology. Additionally, they are able to discriminate between prodromal AD and its advanced phases. About p-tau217, Barthelemy et al. [42] shown that different phosphorylation profiles of soluble tau in CSF and blood plasma in relation to amyloidosis function as a characteristic that separates the stages of AD.

By utilising Förster Resonance Energy Transfer (FRET) to target the proteins p-tau181 and amyloid beta (A*β*-42) and other targets, Balci et al. [43] created a paper-based QD aptasensor for the early detection of AD. Using a Whatman paper with six detecting wells, the sensor integrates hydrophilic and hydrophobic regions—hydrophobic portions made possible by wax. Following their placement in the inlet, blood samples are distributed among six sensing wells that contain QD-aptamer-AuNP complexes. Target proteins cause aptamer conformational changes, which cause CdTe QD fluorescence to be quenched. Two wells are used as references, two target amyloid beta-42, and two target p-tau181. Each well’s fluorescence emission spectrum is captured, demonstrating a linear relationship between protein content and fluorescence quenching. After averaging the values from each pair of wells, the average values of the reference wells and the pairs targeting (A*β*–42) and p-tau181 are compared. In order to aggregate 27 imaging, plasma and CSF measurements of amyloid beta, tau [phosphorylated tau (p-tau), total tau t-tau], neuronal damage and inflammation derived from PET, MRI, immunoassays and mass spectrometry tests were incorporated.

In AD, plasma p-tau 181 was inversely altered, which is probably a sign that neurofibrillary tangles are present in the brain [45]. In line with earlier research, this study also showed a strong correlation between the Aβ42/Aβ40 ratio and p-tau 181 and the CDR and MMSE scores [46, 47]. When it came to differentiating between individuals who were not diagnosed with AD and control participants, the p-tau181 biomarker generally showed the highest specificity and sensitivity. These outcomes closely match those of earlier research. Good diagnostic performance was demonstrated by the biomarkers Aβ40, Aβ42, p-tau181 and total tau (t-tau) [48, 49, 50].

### 5.4. Genetic biomarkers

Genetic biomarkers, especially the presence of the APOE ε4 allele, have been associated with a long history of increased risk for AD. APOE ε4 carriers are more likely to display early amyloid deposition and neurodegeneration. Neuroimaging markers, such as PET and MRI, proved invaluable in monitoring early changes associated with progressive alterations in brain structure and function related to AD.

Meeker et al. [44] employed hierarchical clustering. Measures related to neuropsychology and genetics were also included. Using a random forest approach to feature selection, the best predictors of amyloid PET positive for the whole cohort were found. Additionally, models predicted cognitive decline in both amyloid PET-positive individuals and the cohort as a whole. The findings show that while the number of biomarkers has increased quickly, few of them significantly predict clinical outcomes, and most of them are related to one another. A meaningful framework for understanding the pathobiological change associated with Alzheimer’s disease and information about which biomarkers could be most helpful in clinical treatment and research related to the condition can be obtained by simultaneously examining the whole corpus of known biomarkers.

These biomarkers are promising, but several issues arise from the use of such biomarkers. The significant challenge relates to variability concerning sensitivity and specificity from one biomarker to another across populations and various types of study designs. While Aβ42 and tau have appeared to have high levels of diagnostic accuracy in some cohorts, performance of the biomarkers may vary due to disease progression and the technique(s) of measurement. This review underscores the significance of standardization protocols in the process of evaluating biomarkers to reduce variability and, therefore, enhance the reproducibility of results from studies.

## 6. Conclusion

The systematic review suggests that this broad array of emerging biomarkers with respect to CSF, plasma, and imaging modalities has vast utility in the context of early diagnosis of Alzheimer’s disease. The key biomarkers include Aβ42, tau proteins, neurofilament light chain, and genetic factors, such as APOE ε4, which describe individuals at risk for cognitive decline. However, there are also a few remaining important challenges, including with regard to standardisation of measurement techniques and thereby replication across diverse cohorts. The integrated use of fluid biomarkers with advanced neuroimaging techniques holds promise in new generations of diagnostic frameworks but further research is required to translate these findings into the clinical practises.

## Data Availability

All data produced in the present study are available upon reasonable request to the authors

## 7. Future Directions

Future research on Alzheimer’s Disease (AD) biomarkers should focus on a number of specific areas: multimodal biomarker methods based on fluid, genetic, and neuroimaging data, aimed at more efficient detection at an early stage, especially in asymptomatic subjects; standardization of measurement procedures across studies; population-specific studies in order to disentangle the effects of demographics on the efficacy of the biomarkers; large longitudinal studies intended to validate the diagnostic and prognostic value of new biomarkers over time; and the development of cost-effective, non-invasive screening methods, with special attention on blood-based biomarkers accessible as alternative methods instead of invasive procedures and expensive neuroimaging.

## Funding Statement

**The author(s) received no specific funding for this work**.

## Author Declarations

**I confirm all relevant ethical guidelines have been followed, and any necessary IRB and/or ethics committee approvals have been obtained**.

**The details of the IRB/oversight body that provided approval or exemption for the research described are given below:**

**I confirm that all necessary patient/participant consent has been obtained and the appropriate institutional forms have been archived, and that any patient/participant/sample identifiers included were not known to anyone (e.g., hospital staff, patients or participants themselves) outside the research group so cannot be used to identify individuals**.

**I understand that all clinical trials and any other prospective interventional studies must be registered with an ICMJE-approved registry, such as ClinicalTrials.gov. I confirm that any such study reported in the manuscript has been registered and the trial registration ID is provided (note: if posting a prospective study registered retrospectively, please provide a statement in the trial ID field explaining why the study was not registered in advance)**.

**I have followed all appropriate research reporting guidelines, such as any relevant EQUATOR Network research reporting checklist(s) and other pertinent material, if applicable**.

## Notes

### Competing Interest Statement

The authors have declared no competing interest.

### Funding Statement

This study did not receive any funding

